# Towards reduction in bias in epidemic curves due to outcome misclassification through Bayesian analysis of time-series of laboratory test results: Case study of COVID-19 in Alberta, Canada and Philadelphia, USA

**DOI:** 10.1101/2020.04.08.20057661

**Authors:** Igor Burstyn, Neal D. Goldstein, Paul Gustafson

## Abstract

The aim of our work was to better understand misclassification errors in identification of true cases of COVID-19 and to study the impact of these errors in epidemic curves. We examined publically available time-series data of laboratory tests for SARS-CoV-2 viral infection, the causal agent for COVID-19, to try to explore, using a Bayesian approach, about the sensitivity and specificity of the PCR-based diagnostic test. Data originated from Alberta, Canada (available on 3/28/2020) and city of Philadelphia, USA (available on 3/31/2020). Our analysis revealed that the data were compatible with near-perfect specificity but it was challenging to gain information about sensitivity (prior and posterior largely overlapped). We applied these insights to uncertainty/bias analysis of epidemic curves into jurisdictions under the assumptions of both improving and degrading sensitivity. If the sensitivity improved from 60 to 95%, the observed and adjusted epidemic curves likely fall within the 95% confidence intervals of the observed counts. However, bias in the shape and peak of the epidemic curves can be pronounced, if sensitivity either degrades or remains poor in the 60-70% range. In the extreme scenario, hundreds of undiagnosed cases, even among tested, are possible, potentially leading to further unchecked contagion should these cases not self-isolate. The best way to better understand bias in the epidemic curves of COVID-19 due to errors in testing is to empirically evaluate misclassification of diagnosis in clinical settings and apply this knowledge to adjustment of epidemic curves, a task for which the Bayesian method we presented is well-suited.

## Introduction

It is well known that outcome misclassification can bias epidemiologic results, yet is infrequently quantified and adjusted for in results. In the context of infectious disease outbreaks, such as during the COVID-19 pandemic of 2019-20, false positive diagnoses may lead to a waste of limited resources, such as testing kits, hospital beds, and absence of the healthcare workforce. On the other hand, false negative diagnoses contribute to uncontrolled spread of contagion, should these cases not self-isolate. In an ongoing epidemic, where test sensitivity (Sn) and specificity (Sp) of case ascertainment are fixed, prevalence of the outcome (infection), determines whether false positives or negatives dominate. For COVID-19, Goldstein & Burstyn (2020) show that suboptimal test Sn despite excellent Sp results in an overestimation of cases in the early stages of an outbreak, and substantial underestimation of cases as prevalence increases to levels seen at the time of writing. However, understanding the true scope of the pandemic depends on precise insights into accuracy of laboratory tests used for case confirmation. Undiagnosed cases are of particular concern; they arise from untested persons who may or may not have symptoms (under-ascertainment) and from errors in testing among those selected for the test (inconclusive results). We focus on misclassified patients only due to errors in tests that were performed: according to the World Health Organization’s case definition, these may be deemed *probable* cases when a test result was inconclusive (WHO, 2020). Presently, the accuracy of testing for SARS-CoV-2 viral infection, the causal agent for COVID-19, is unknown in Canada and the USA, but globally it is reported that Sp exceeds Sn (COVID-19 Science Report: Diagnostics 2020, Fang et al. 2020; Ai et al. 2020).

In a typical scenario, clinical and laboratory validation studies are needed to fully quantify the performance of a diagnostic assay (measured through Sn and Sp). However, during a pandemic, limited resources are likely to be allocated to testing and managing patients, rather than performing the validation work. After all, imperfect testing can still shed a crude light on the scope of the public health emergency. Indeed, counts of observed positive and negative tests can be informative about Sn and Sp, because certain combinations of these parameters are far more likely to be compatible with data and reasonable assertions about true positive tests. In general, more severe cases of disease are expected at the onset of an outbreak (and reflected in tested samples as strong clinical suspicion for the test produces higher likelihood of having the disease) but the overall prevalence in the population would remain low. Then, as the outbreak progresses with more public awareness and consequently both symptomatic and asymptomatic people being tested, the overall prevalence of disease is expected to rapidly increase while the severity of the disease at a population level is tempered. It is reasonable to expect, as was indeed reported anecdotally early in the COVID-19 outbreak, for laboratory tests to be inaccurate, because the virus itself and its unique identifying features exploited in the test are themselves uncertain, and laboratory procedures can contain errors ahead of standardization and regulatory approval. Again, anecdotally, Sn was supposed to be worse than Sp, which is congruent with reports of early diagnostic tests from China (Fang et al. 2020; Ai et al. 2020), with both Sn and Sp improving as the laboratories around the world rushed to perfect testing (Konrad et al. 2020; US FDA 2020; <u>Corman</u> et al. 2020) to approach performance in tests for similar viruses (e.g. Binsaeed et al. 2011; Merckx et al., 2017). Using publicly available time-series data of laboratory testing results for SARS-CoV-2 and our prior knowledge of infectious disease outbreaks, we may be able to gain insights into the true accuracy of the diagnostic assay.

Thus, we pursued two specific aims: (a) to develop a Bayesian method to attempt to learn from publicly available time-series of COVID-19 testing about Sn and Sp of the laboratory tests and (b) to conduct a Monte Carlo (probabilistic) sensitivity analysis of the impact of the plausible extent of this misclassification on bias in epidemic curves.

## Methods

### Sharing

Data and methods can be accessed at https://github.com/paulgstf/misclass-covid-19-testing; data are also displayed in Appendix A.

### Data

We digitized data released by the Canadian province of Alberta on 3/28/2020 from their “Figure 6: People tested for COVID-19 in Alberta by day” on https://covid19stats.alberta.ca/ under “Laboratory testing” tab. Samples (e.g., nasopharyngeal (NP) swab; bronchial wash) undergo nucleic acid testing (NAT) that use primers/probes targeting the E (envelope protein) (Corman et al. 2020) and RdRp (RNA-dependent RNA polymerase) (qualitative detection method developed at ProvLab of Alberta) genes of the COVID-19 virus. The data was digitized as shown in Table A1 of Appendix A. The relevant data notes are reproduced in full here:

“Data sources: The Provincial Surveillance Information system (PSI) is a laboratory surveillance system which receives positive results for all Notifiable Diseases and diseases under laboratory surveillance from Alberta Precision Labs (APL). The system also receives negative results for a subset of organisms such as COVID-19. … Disclaimer: The content and format of this report are subject to change. Cases are under investigation and numbers may fluctuate as cases are resolved. Data included in the interactive data application are up-to-date as of midday of the date of posting.”

Data from the city of Philadelphia were obtained from https://www.phila.gov/programs/coronavirus-disease-2019-covid-19/the-citys-response/monitoring-and-testing/ on 03/31/2020. It was indicated that “test results might take several days to process.” Most testing is PCR-based with samples collected from an NP swab, performed at one of the three labs (State Public Health, Labcorps, Quest). In addition, some hospitals perform this test using ‘in-house’ PCR methods. There is a perception (but no empiric data available to us) that Sn is around 0.7 and there are reports of false negatives based on clinical features of patients consistent with COVID-19 disease. Issues arise from problems with specimen collection and timing of the collection, in addition to test performance characteristics. The data were digitized as shown in Table A2 in Appendix A.

### Bayesian method to infer test sensitivity and specificity

A brief description of the modeling strategy follows here, with full details in Appendix B. Both prevalence of infection in the testing pool and test sensitivity are modeled as piecewise-linear on a small number of adjacent time intervals (here: days; four intervals of equal width, in both examples), with the interval endpoints referred to as “knots” (hence there are five knots, in both examples). The prior distribution for prevalence is constructed by specifying lower and upper bounds for prevalence at each knot, with a uniform distribution in between these bounds. The prior distribution of sensitivity is constructed similarly, but with a modification to encourage more smoothness in the variation over time (see Appendix B for full details). The test specificity is considered constant over time, with a uniform prior distribution between specified lower and upper bounds.

With the above specification, a posterior distribution ensues for all the unknown parameters and latent variables given the observed data, i.e., given the daily counts of negative and positive test results. This distribution describes knowledge of prevalence, sensitivity, specificity, and the time-series of the latent Y_t_, the number of truly positive among those tested on the t-th day. Thus, we learn the posterior distribution of the Y_t_ time-series, giving an adjusted series for the number of true positives in the testing pool, along with an indication of uncertainty.

As discussed at more length in Appendix B, this model formulation neither rules in, nor rules out, learning about test sensitivity and specificity from the reported data. Particularly in a high specificity regime, the problem of separating out infection prevalence and test sensitivity is mathematically challenging. The data directly inform only the product of prevalence and sensitivity. Trying to separate the two can be regarded statistically as an “unidentified” problem (while mathematicians might speak of an ill-posed inverse problem, or engineers might refer to a “blind source separation” challenge). However, some circumstances might be more amenable to some degree of separation. In particular, with piecewise-linear structure for sensitivity and prevalence, strong quadratic patterns in the observed data, if present, could be particularly helpful in guiding separation. On the other hand, if little or no separation can be achieved, the analysis will naturally revert back to a sensitivity analysis, with the *a priori* uncertainty about test sensitivity and infection prevalence being acknowledged.

Some of the more reliable PCR-based assays can achieve near-perfect Sp and Sn of around 0.95 (Konrad et al. 2020; US FDA 2020; Corman et al. 2020, COVID-19 Science Report: Diagnostics 2020). However, early in the COVID-19 outbreak problems with the sensitivity of the diagnostic test were widely reported owing to specimen collection and reagent preparation, but not quantified. Based on these reports, we posited a lower bound on prior Sn of 0.6 and an upper bound of 0.9. We expected Sp to be high and selected a time-invariant prior uniform bounded by 0.95 and 1. Prevalence of test-positive samples likely changed over time as well -- for example due to prioritization of testing based on age, occupation, and morbidity (CDC, 2020) -- but it is difficult to quantify what this would be, because it is different from population prevalence of infected yielded by a random sample (governed by known population dynamics models). Consequently, we adopt a flexible data-driven approach by allowing prevalence to change within broad ranges (see Appendix B)

### Monte Carlo (probabilistic) uncertainty/bias analysis of epidemic curves

We next examined how much more we could have learnt from epidemic curves if we knew sensitivity of laboratory testing. To do so, we applied insights into the plausible extent of sensitivity and specificity to re-calculate epidemic curves for COVID-19 in Alberta, Canada. Data on observed counts versus presumed incident dates (“date reported to Alberta Health”) was obtained on 3/28/2020 from their “Figure 3: COVID-19 cases in Alberta by day” posted on https://covid19stats.alberta.ca/ under “Case counts” tab. The count of cases is shown in Table A1 as C_t_^*^ and they are matched to dates t (same as dates of laboratory tests). We also repeated these calculations with data available for City of Philadelphia, under a strong assumption that date of tests is the same as date of onset, i.e. Y_t_^*^ = C_t_^*^ We removed March 30-31, 2020, counts because of a reported delay of several days in laboratory tests.

For each observed count of incident cases C_t_^*^, we estimated true counts 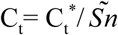 under the assumption that specificity is indistinguishable from perfect. We considered a situation of no time trend in line with above findings, as well as sensitivity either improving (realistic best case), or degrading (pessimistic worst case). We simulated various values of 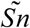 using *Beta* distribution ranging in means from 0.60 to 0.95, with a fixed standard deviation of 0.05 (parameters set using https://www.desmos.com/calculator/kx83qio7yl). It is apparent that epidemic curves generated in this manner will have higher counts than the observed curves, and our main interest is to illustrate how much the underestimation can bias the depiction. Our uncertainty/bias analysis only reflects systematic errors for illustrative purposes and under the common assumptions (and experience) that they dwarf random errors. Computing code in R (R Core Team, 2019) for the uncertainty analysis is in Appendix C.

## Results

### Inference about sensitivity and specificity

In both jurisdictions, there is evidence of non-linearity in the observed proportion of positive tests (Figure A), justifying our flexible approach to variation of sensitivity and prevalence that can be manifest by quadratic pattern in observed prevalence between knots. The data in both jurisdictions is consistent with the hypothesis that *the number of truly infected is being under-estimated*, even though observed counts tend to fall within 95% credible intervals of posterior distribution of the counts of true positive tests (Figure B). The under-diagnosis is more pronounced when there are both more positive cases and the prevalence of positive tests is higher, i.e. in Philadelphia related to Alberta. In Philadelphia, the posterior of prevalence was between 5 and 24% (100’s of positive tests a day in late March) but in Alberta, the median of the posterior of prevalence was under 3% (30 to 50 positive tests a day in late March). This is not surprising because the number of false negatives is proportional to observed cases for the same sensitivity. The specificity appears to be high enough for the observed prevalence to produce negligible numbers of false positives, with false negatives dominating. There was clear evidence of shift in posterior distribution of *specificity* from uniform to the one centered values of >0.98 (Figure C). In Alberta, posterior distribution of Sp was centered on 0.997 (95% credible interval (CrI): 0.993, 0.99995), and in Philadelphia it had a posterior median of 0.984 (95%CrI: 0.954, 0.999). Our analysis indicates that under our models there is little evidence in time-series of laboratory tests about either the time trend or magnitude of *sensitivity* of laboratory tests in either jurisdiction (Figure D). Posterior distributions are indistinguishable from the priors, such that we are still left with an impression that sensitivity of COVID-19 tests can be anywhere between 0.6 and 0.9, centered around 0.75. One can speculate on the departure of the posterior distribution from uniform prior given that the posterior appears concentrated somewhat around the prior mean of 0.75 (more lines in Figure D near the mean than the dotted edges that bound the prior). However, any such signal is weak and there is no evidence of a time-trend that was allowed by the model.

**Figure A:**
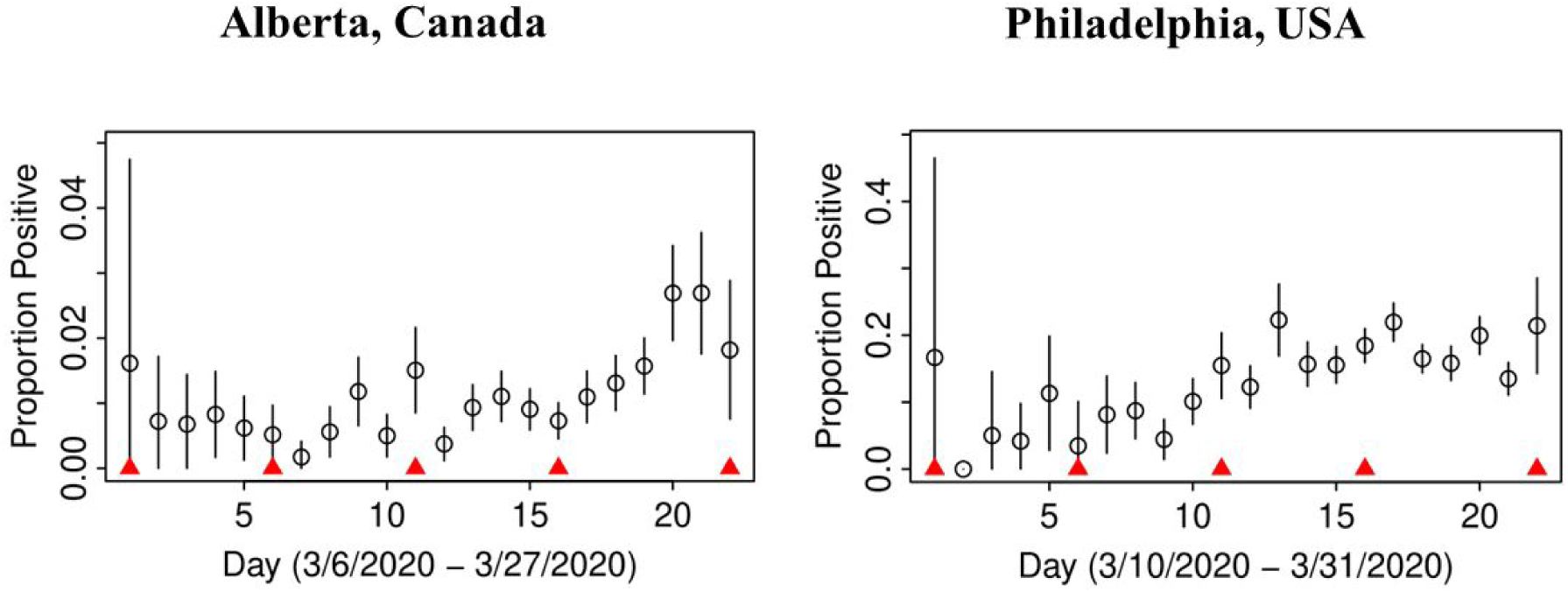
Proportion of observed positive tests in time with 95% confidence intervals; knots between which sensitivity and true prevalence were allowed to follow linear trends are indicated by red triangles.

**Figure B:**
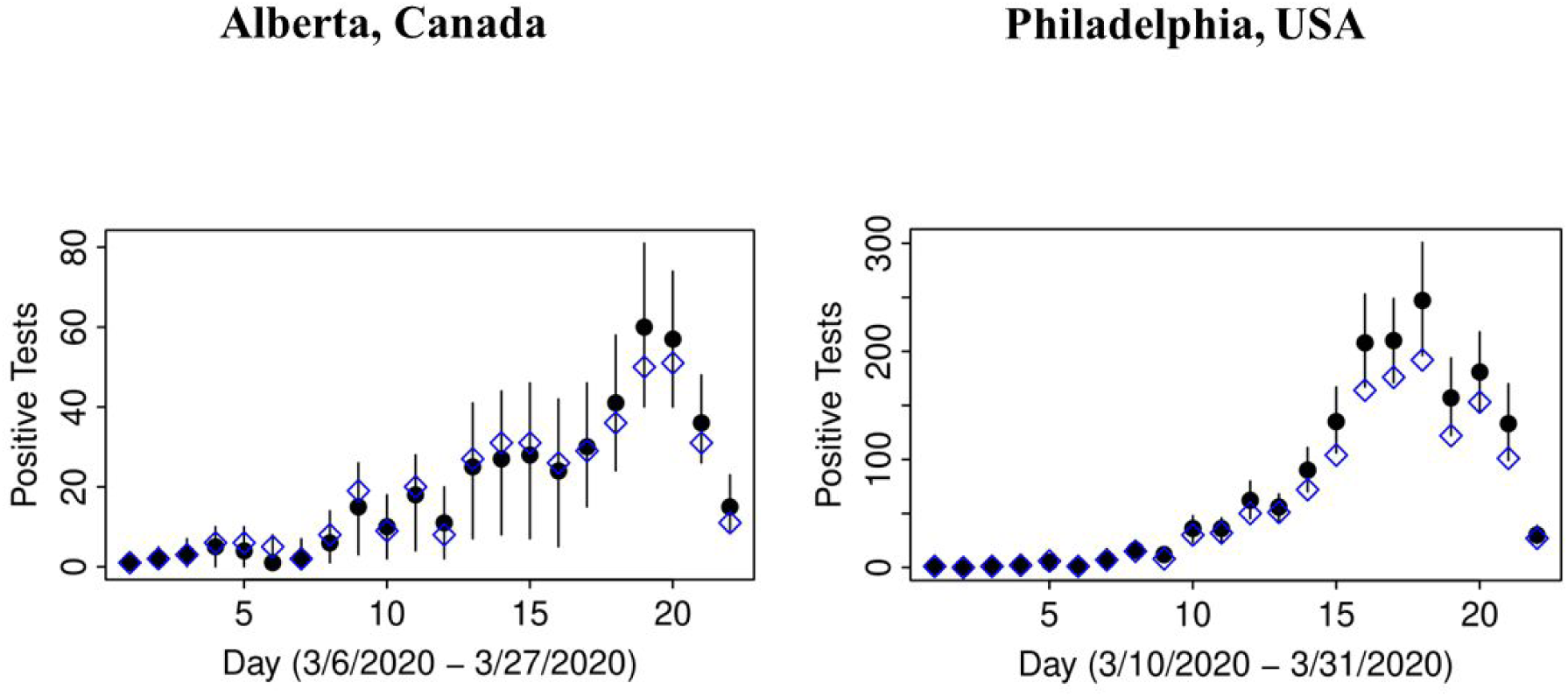
Observed count of positive tests for COVID-19 (open diamond) versus posterior distributions of counts, adjusted for misclassification (solid circles as means and lines representing 95% credible intervals)

**Figure C:**
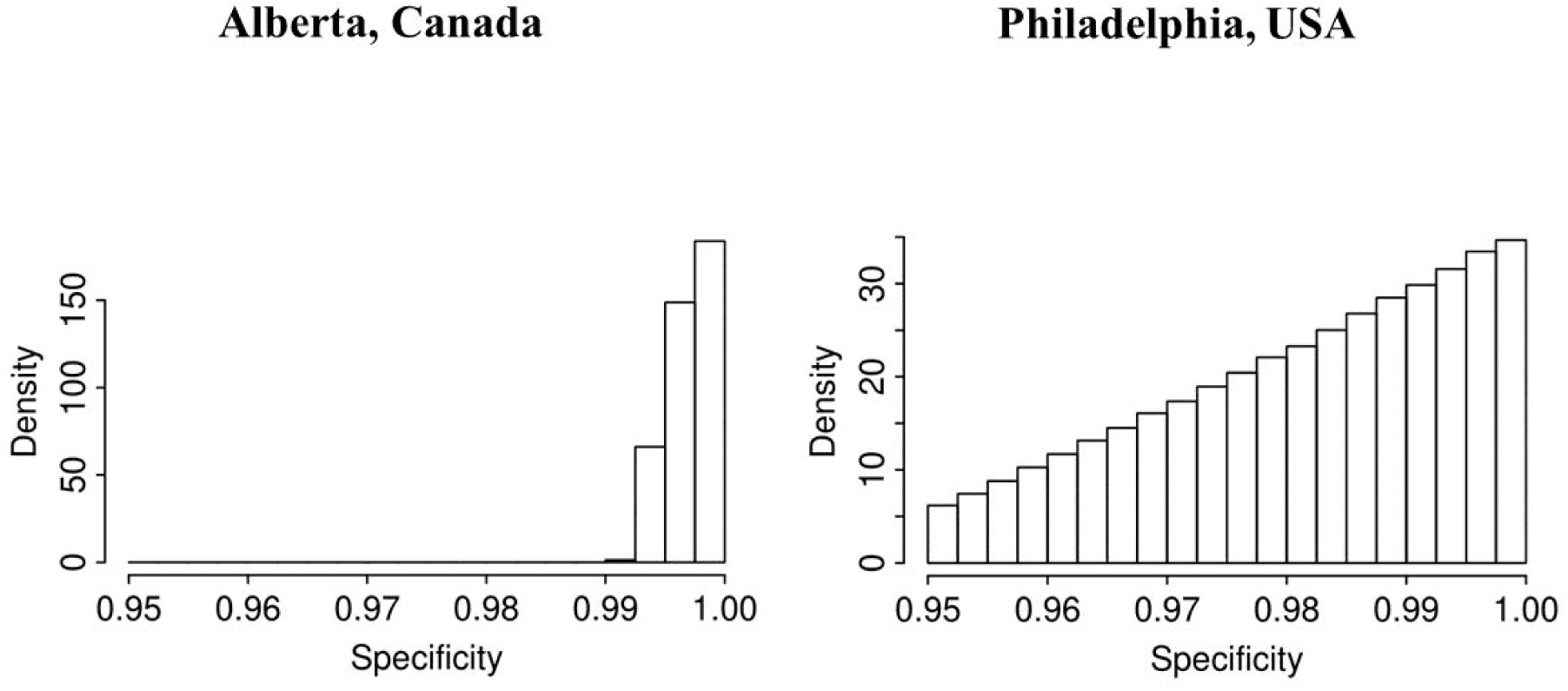
Histograms of posterior distributions of specificity of laboratory test for COVID-19

**Figure D:**
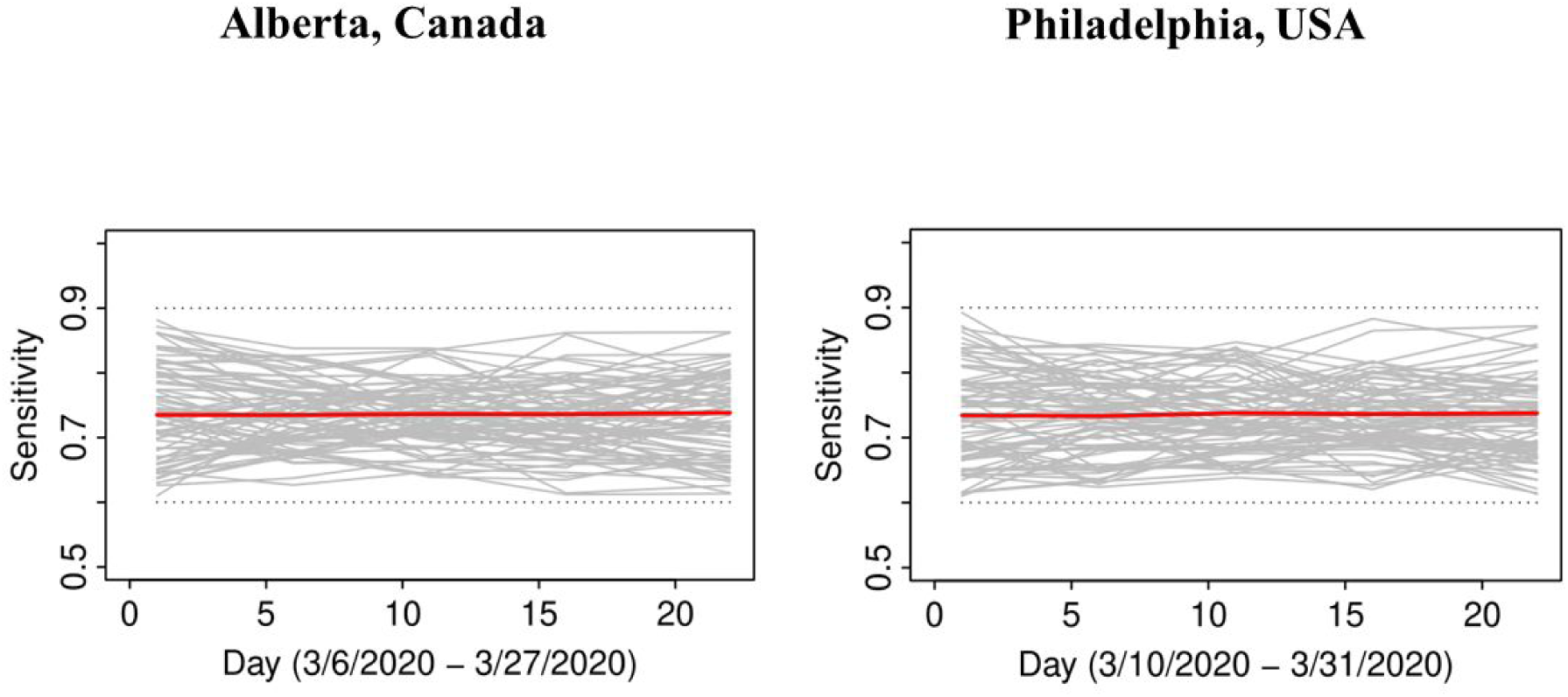
Posterior distributions of sensitivity of of laboratory test for COVID-19 in time (grey), posterior mean (red), and boundaries of priors (dotted)

### Uncertainty in epidemic curves due to imperfect testing: Alberta, Canada

The left-hand side panel of Figure E presents an impact on the epidemic curve of degrading sensitivity that is constant in time. As expected, when misclassification errors increase, uncertainty about epidemic curves also increases. There is an under-estimation of incident cases that is more apparent later in the epidemic when the numbers rise. The right-hand panel of Figure E indicates how, as expected, if sensitivity improves over time (green lines), then the true epidemic curve is expected to be flatter than the observed. It also appears that observed and true curves may well fall within the range of 95% confidence intervals around the observed counts (blue lines). If sensitivity decreases over time (brown lines), then the true epidemic curve is expected to be steeper than the observed. In either scenario, there can be an under-counting of cases by nearly a factor of two, most apparent as the incidence grows, such that on day March 24, 2020 (t=19), there may have been almost 120 cases vs. 62 observed. This is alarming, because misdiagnosed patients can spread infection if they have not self-isolated (perhaps a negative test results provided a false sense of security) and it is impossible to know who they are among thousands of symptomatic persons tested around that time per day (Table A1).

**Figure E:**
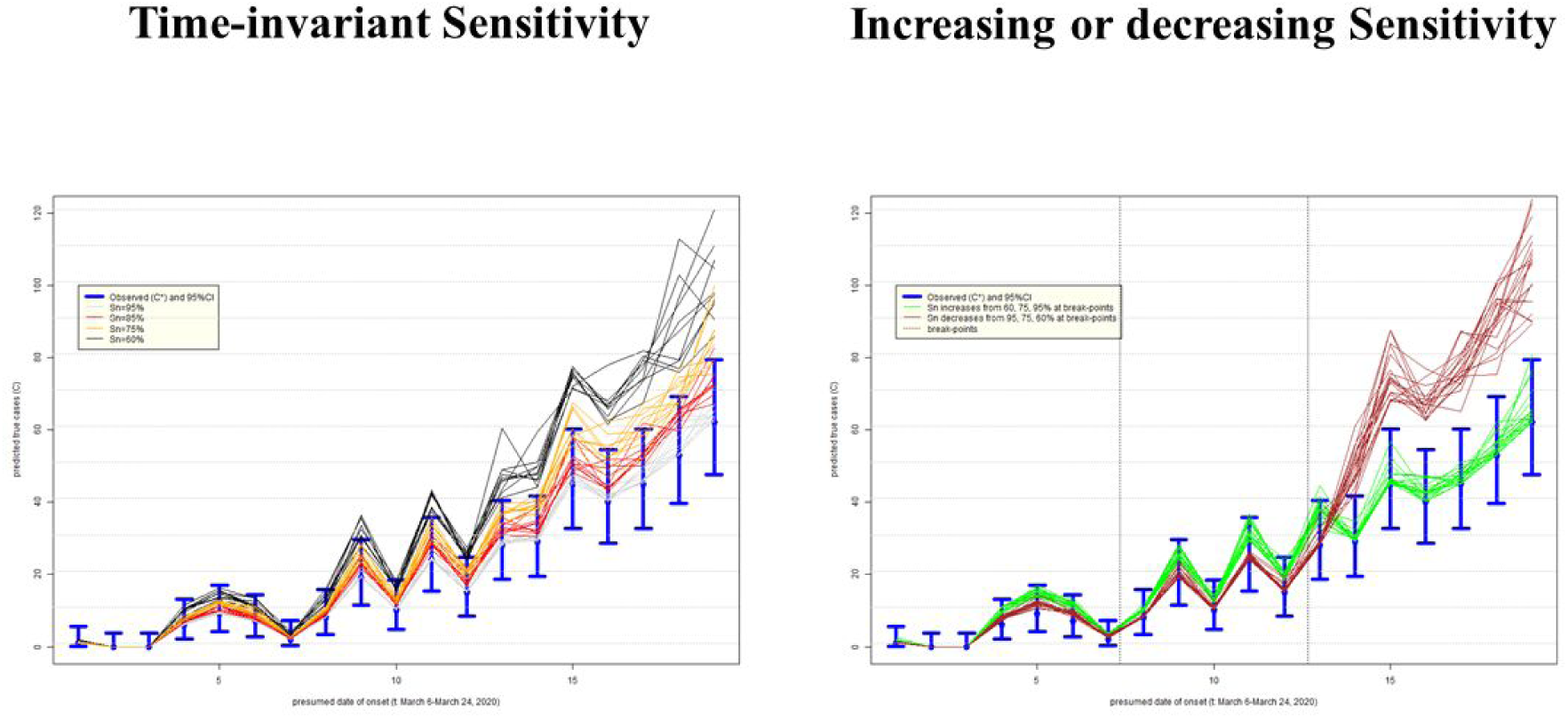
Uncertainty in the epidemic curve of COVID-19 on March 28, 2020 in Alberta, Canada, due to imperfect sensitivity (Sn) with standard deviation 5%; assumes specificity 100%

### Uncertainty in epidemic curves due to imperfect testing: Philadelphia, USA

The left-hand panel of Figure F presents an impact on the epidemic curve of degrading Sn that is constant in time. As in Alberta, when misclassification errors increase, uncertainty about epidemic curves also increases. It is also apparent that the shape of the epidemic curve, especially when counts are high, can be far steeper than that inferred assuming perfect testing. The right-hand panel of Figure F indicates that if sensitivity improves over time (green lines), then the true epidemic curve is expected to be practically indistinguishable from the observed one in Philadelphia: e.g. it is within random variation of observed counts represented by 95% confidence intervals (blue lines). This is comforting, because this seems to be the most plausible scenario of improvement in time in quality of testing (identification of truly infected). However, if sensitivity decreases over time (brown lines), then the under-counting of cases by the hundreds in late March 2020 cannot be ruled out. We again have the same concern as for Alberta: misdiagnosed patients can spread infection unimpeded and it is impossible to know who they are among the hundreds of symptomatic persons tested in late March 2020 (Table A2).

**Figure F:**
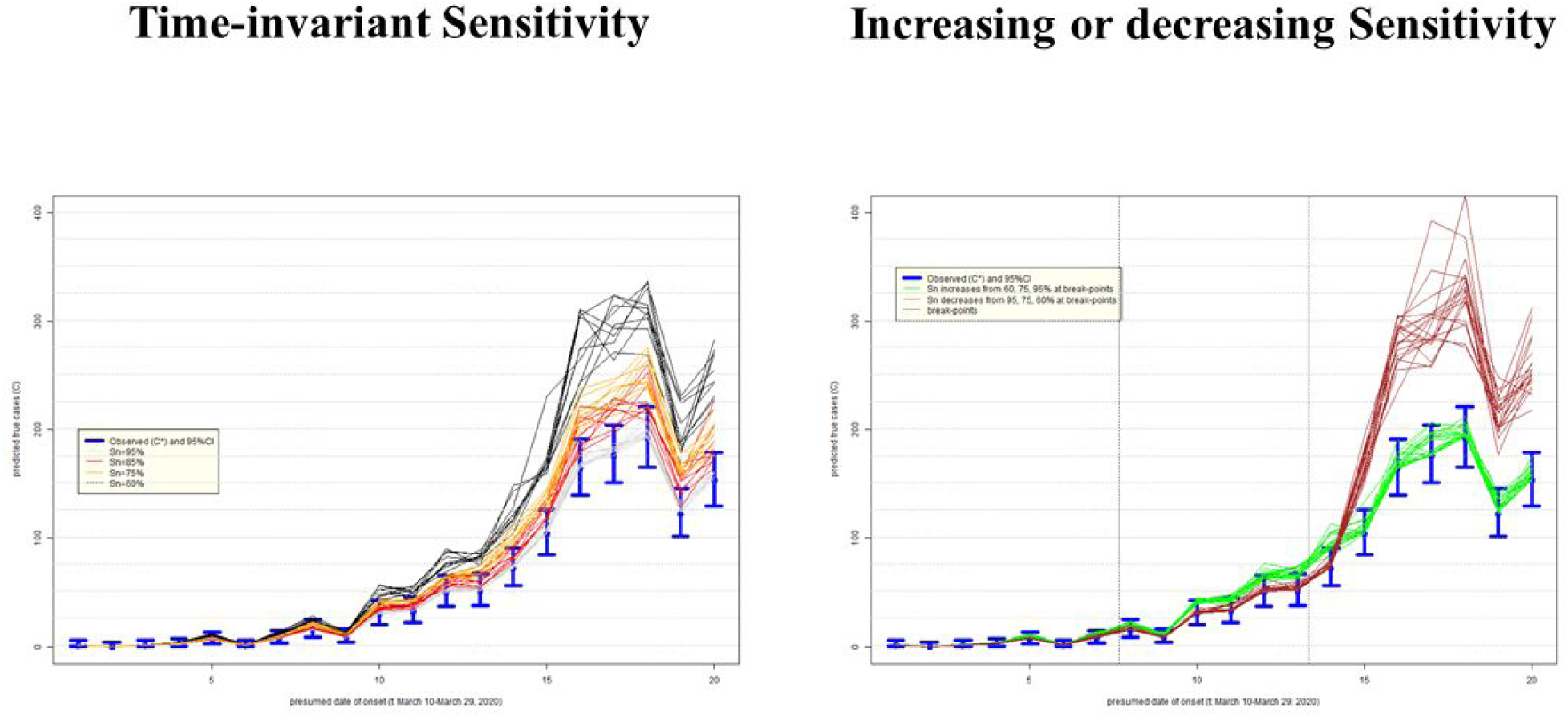
Uncertainty in the epidemic curve of COVID-19 on March 31, 2020 in Philadelphia, USA, due to imperfect sensitivity (Sn) with standard deviation 5%; assumes specificity 100%

In all examined scenarios, in both Alberta and Philadelphia, the lack of sensitivity in testing seems to matter far less when the observed counts are low early in the epidemic. The gap between observed and adjusted counts grows as the number of observed cases increases. This reinforces the importance of early testing, at least with respect to describing the time-course of the epidemic, even with imperfect technology (Goldstein & Burstyn, 2020).

## Discussion

Given the current uncertainty in the accuracy of the SARS-CoV-2 diagnostic assays, we tried to learn about sensitivity and specificity using the time-series of laboratory tests and time trends in time test results. Although we are confident that *typical specificity exceeds 0*.*98*, there is *very little learning about sensitivity* from prior to posterior. However, it is important to not generalize this lack of learning about sensitivity, because it can occur when stronger priors on prevalence are justified and/or when there are more pronounced trends in prevalence of positive tests. We therefore encourage every jurisdiction with suitable data to attempt to gain insights into accuracy of tests using our method: now that the method to do so exists, it is simpler and cheaper than laboratory and clinical validation studies. However, validation studies, with approaches like the one illustrated in Burstyn et al. (2009), are still the most reliable means of determining accuracy of a diagnostic test.

Knowing sensitivity and specificity is important as demonstrated in uncertainty/bias analysis of impact on epidemic curves under some optimistic assumptions of near-perfect specificity and reasonable range of sensitivity. The observed epidemic curves may bias estimates of the effective reproduction number (R_e_) and magnitude of the epidemic (peak) in unpredictable directions. This may also have implications for understanding the proportion of the population non-susceptible to COVID-19. As researchers attempt to develop pharmaceutical prophylaxis (i.e., a vaccine) combined with a greater number of people recovering from SARS-CoV-2 infection, having insight into the herd threshold will be important for resolving current and future outbreaks. Calculations such as the basic and effective reproductive number, and the herd threshold depend upon the accuracy of surveillance data described in the epidemic curves.

Limitations of our approach include the dynamic nature of data that changes daily and may not be perfectly aligned in time due to batch testing. There are some discrepancies in the data that should be resolved in time, like fewer cases tested positive than there are in epidemic curve in Alberta, *but the urgency of the current situation justifies doing our best with what we have now*. We also make some strong *ad hoc* assumptions about breakpoints in segmented regression of time-trends in sensitivity and prevalence, further assuming that the same breakpoints are suitable for trends in both parameters. Although not as much of an issue based on our analysis, we do need to consider imperfect specificity, creating false positives, albeint nowhere near the magnitude of false negatives in the middle of an outbreak. This results in wasted resources. In ideal circumstances we employ a two stage test: a highly sensitive serological assay, that if positive triggers a PCR-based assay. Thus we would require two samples per individual: blood and NP swab. Two-stage tests would resolve a lot of uncertainty and speculation over a single PCR test combined with signs and symptoms. Indeed, this is the model used for diagnosis of other infectious diseases, such as HIV and Hepatitis C. Our work also only focuses on validity of laboratory tests, not sensitivity and specificity of the entire process of identification of cases that involves selection for testing via a procedure that is designed to induce systemic biases relative to the population.

## Conclusions

We conclude that it is of paramount importance to validate laboratory tests and to share this knowledge, especially as the epidemic matures into its full force. Insights into ascertainment bias by which people are selected for tests and are then used to estimate epidemic curves are likewise important to obtain and quantify. Quantification of these sources of misclassification and bias can lead to adjusted analyses of epidemic curves that can help make more appropriate public health policies.

## Data Availability

Data and methods can be accessed at https://github.com/paulgstf/misclass-covid-19-testing, as well as Appendix C.

## Acknowledgements

The authors thank Isaac R. Burstyn and Marguerite R. Burstyn for their diligence in perfect extraction of Alberta data from the on-line charts. IB also thanks James L.B. Burstyn for not teething every night during preparation of this manuscript. Nicola Cherry and Esther Chernak earned our gratitude by assisting with insights on testing in Alberta and Philadelphia, respectively. Research reported in this publication was partially supported by the National Institute of Allergy And Infectious Diseases of the National Institutes of Health under Award Number K01AI143356 (to NDG).

## Appendix A: Data used in the manuscript

**Table A1:** Timeline of counts of positive and negative tests for COVID-19 and cases per onset date in Alberta, Canada on 3/28/2020

| Lab report date /date of onset for incident case (t) | test positive ($Y_t^*$)# | test negative ($n_t - Y_t^*$) | Observed incident cases of COVID-19 ($C_t^*$)# |
| --- | --- | --- | --- |
| 2020-03-06 | 1 | 61 | 1 |
| 2020-03-07 | 2 | 275 | 0 |
| 2020-03-08 | 3 | 440 | 0 |
| 2020-03-09 | 6 | 718 | 6 |
| 2020-03-10 | 6 | 966 | 9 |
| 2020-03-11 | 5 | 961 | 7 |
| 2020-03-12 | 2 | 1144 | 2 |
| 2020-03-13 | 8 | 1419 | 8 |
| 2020-03-14 | 19 | 1591 | 19 |
| 2020-03-15 | 9 | 1784 | 10 |
| 2020-03-16 | 20 | 1306 | 24 |
| 2020-03-17 | 8 | 2137 | 15 |
| 2020-03-18 | 27 | 2860 | 28 |
| 2020-03-19 | 31 | 2774 | 29 |
| 2020-03-20 | 31 | 3392 | 45 |
| 2020-03-21 | 26 | 3539 | 40 |
| 2020-03-22 | 29 | 2613 | 45 |
| 2020-03-23 | 36 | 2713 | 53 |
| 2020-03-24 | 50 | 3131 | 62 |
| 2020-03-25 | 51 | 1842 | 77 |
| 2020-03-26 | 31 | 1121 | 48 |
| 2020-03-27 | 11 | 593 | 14 |

#the number of persons who tested positive at time $t$ is not the same as number with persons with onset of symptoms at time $t$ (used in epidemic curve); the lack of agreement two counts and their totals is due to lag in reporting and investigation of date of onset, a situation that is expected in the middle of epidemic at the time of compilation of these data

**Table A2:** Timeline of positive and negative tests for COVID-19 in Philadelphia, USA; obtained from figures published by City of Philadelphia on 3/31/2020

| date of test report (t) | test positive ($Y_t^*$) | test negative ($n_t - Y_t^*$) |
| --- | --- | --- |
| 3/10/2020 | 1 | 5 |
| 3/11/2020 | 0 | 13 |
| 3/12/2020 | 1 | 19 |
| 3/13/2020 | 2 | 46 |
| 3/14/2020 | 6 | 47 |
| 3/15/2020 | 1 | 28 |
| 3/16/2020 | 7 | 79 |
| 3/17/2020 | 15 | 157 |
| 3/18/2020 | 8 | 173 |
| 3/19/2020 | 30 | 267 |
| 3/20/2020 | 32 | 175 |
| 3/21/2020 | 50 | 358 |
| 3/22/2020 | 51 | 178 |
| 3/23/2020 | 72 | 387 |
| 3/24/2020 | 104 | 564 |
| 3/25/2020 | 164 | 725 |
| 3/26/2020 | 176 | 626 |
| 3/27/2020 | 192 | 973 |
| 3/28/2020 | 122 | 650 |
| 3/29/2020 | 153 | 614 |
| 3/30/2020 | 101 | 647 |
| 3/31/2020 | 27 | 99 |

## Appendix B: Statistical Modeling Details

Let r_k_ be the infection prevalence in the testing pool on the day corresponding to the k-th of K knots, and let Sn_k_ be the test sensitivity achieved within this pool. Without ambiguity, when we index with t to indicate the t-th of T days (as opposed to k for the k-th of K knots), r_t_ and Sn_t_ to be the prevalence and sensitivity achieved via linear interpolation from the straddling knots. Let Sp be the test specificity, assumed to be constant in time.

A prior for prevalence is specified as a distribution for r_1:K_ with independent uniform components. The left and right endpoints for each component then comprise 2K hyperparameters, though in our applications we reduce this to a single hyperparameter, by specifying a common upper-bound while setting zero as a common lower-bound.

A prior for sensitivity is constructed by taking Sn_k_ = (1-w)Sn_A,k_ + w Sn_B,k_. Here the prior specification for Sn_A:1:K_ is via independent uniform components with the hyperparameters being the corresponding 2K endpoints. The second component is perfectly linear, so that only (Sn_B,1_, Sn_B,K_) are stochastic, taken as independently and uniformly distributed using the same endpoints as for the corresponding components of Sn_A_. Thus, when w=0 the prior for Sn has the same structure (but different hyperparameters) as the prior for r. As w increases, however, the time trajectories for Sn become smoother. We complete the specification by assigning a prior to w itself, namely w ∼ Unif(0.5, 0.9). Thus, we do not arbitrarily dictate the trajectory smoothness *a priori*. Note also that in our applications we use common lower and upper bounds for Sn_1:K_, so that only two hyperparameters are needed.

Having defined a prior for r, Sn, and Sp, we now specify a model for the observed number of positive tests on the t-th day, 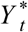, out of n tests. We build this up from a factorization of the form

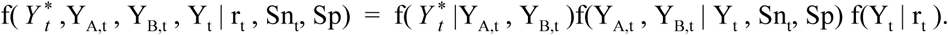

Here Y_t_ ∼ Bin(n_t_, r_t_) is the true number of day t positives, which is ultimately the parameter of interest. Whereas t-th observed count decomposes 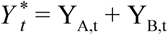, where Y_A,t_ ∼ Bin(Y_t_, Sn_t_) is the portion of the observed count arising as true positives, while independently Y_B,t_ ∼ Bin(n_t_ - Y_t_, 1-Sp) is the portion arising as false positives.

The above comprises a complete stochastic generative model. Hence, Markov chain Monte Carlo sampling from the joint distribution of all latent quantities given all observed quantities is feasible. The latents include w, Sn_A:1:K_, (Sn_B,1_, Sn_B,K_), Sp, r_1:K,_ Y_1:T,_ Y_A,1:T_, and Y_B,1:T_. The observed data are the two series: 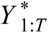 and n_1:T_.

To elaborate on the underidentified nature of the statistical problem, it is simplest to consider the case of Sp=1, in which case the observed data are governed by 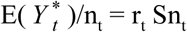 so that indeed asking the data to completely distinguish Sn_t_ from r_t_ is impossible, at least in the absence of prior information. If the piecewise-linear assumptions on Sn_t_ and r_t_ are valid, however, then one might anticipate some partial information coming forward. Consider a single interval between two knots. If the data signal on this interval happens to be highly linear, then we have a severe paucity of information, as two coefficients describing the composite signal are being asked to inform about the four coefficients describing the two constituent signals, albeit with help from the prior bounds on the constituent signals. On the other hand, if the data signal happens to be quadratic, then we still face an underidentified situation, but at least now there are three pieces of information from the composite signal to partially inform the four descriptors on the constituent quantities.

## Appendix C: R code for Figures E and F (code to produce panel as .png files is omitted)

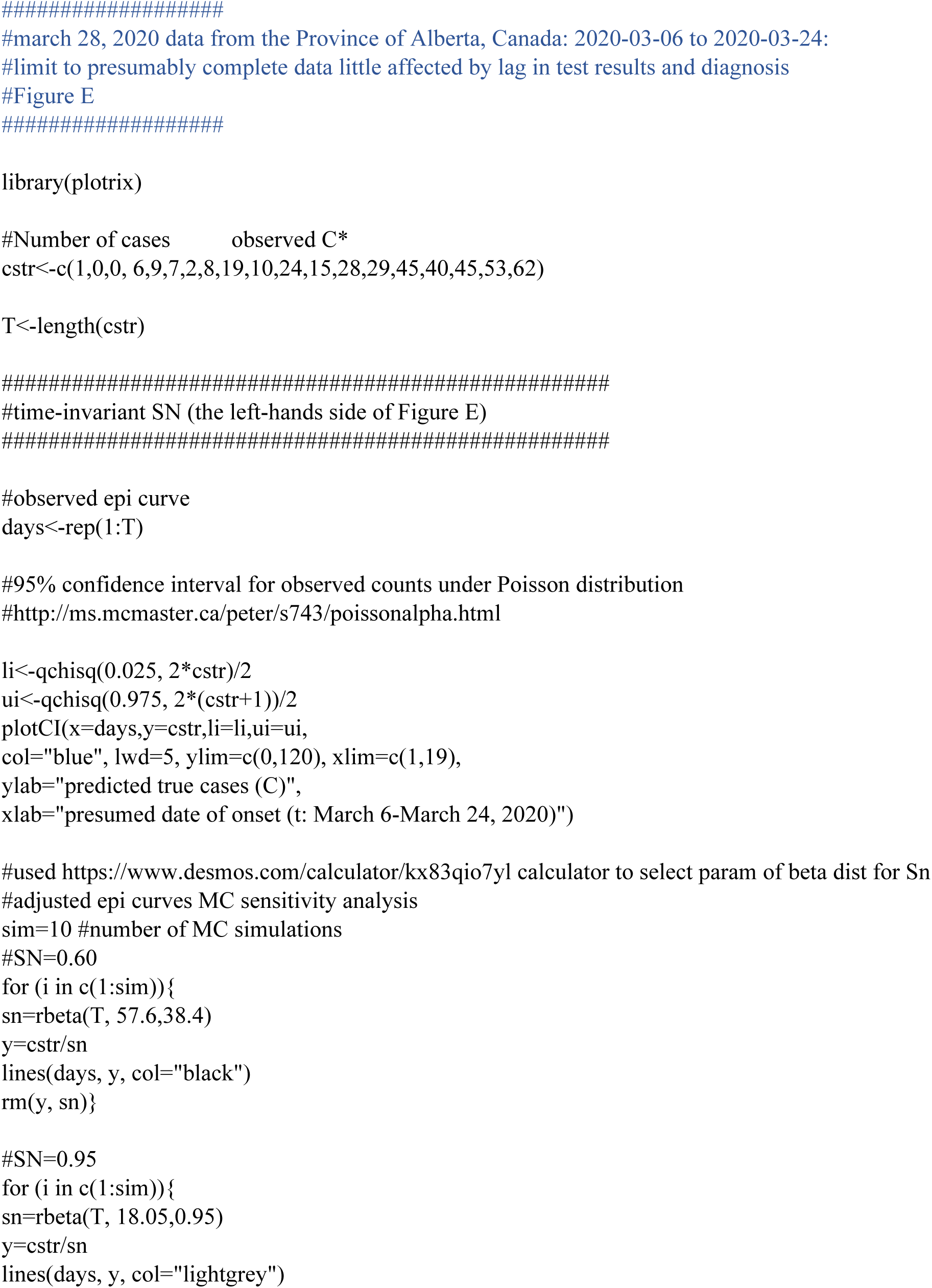
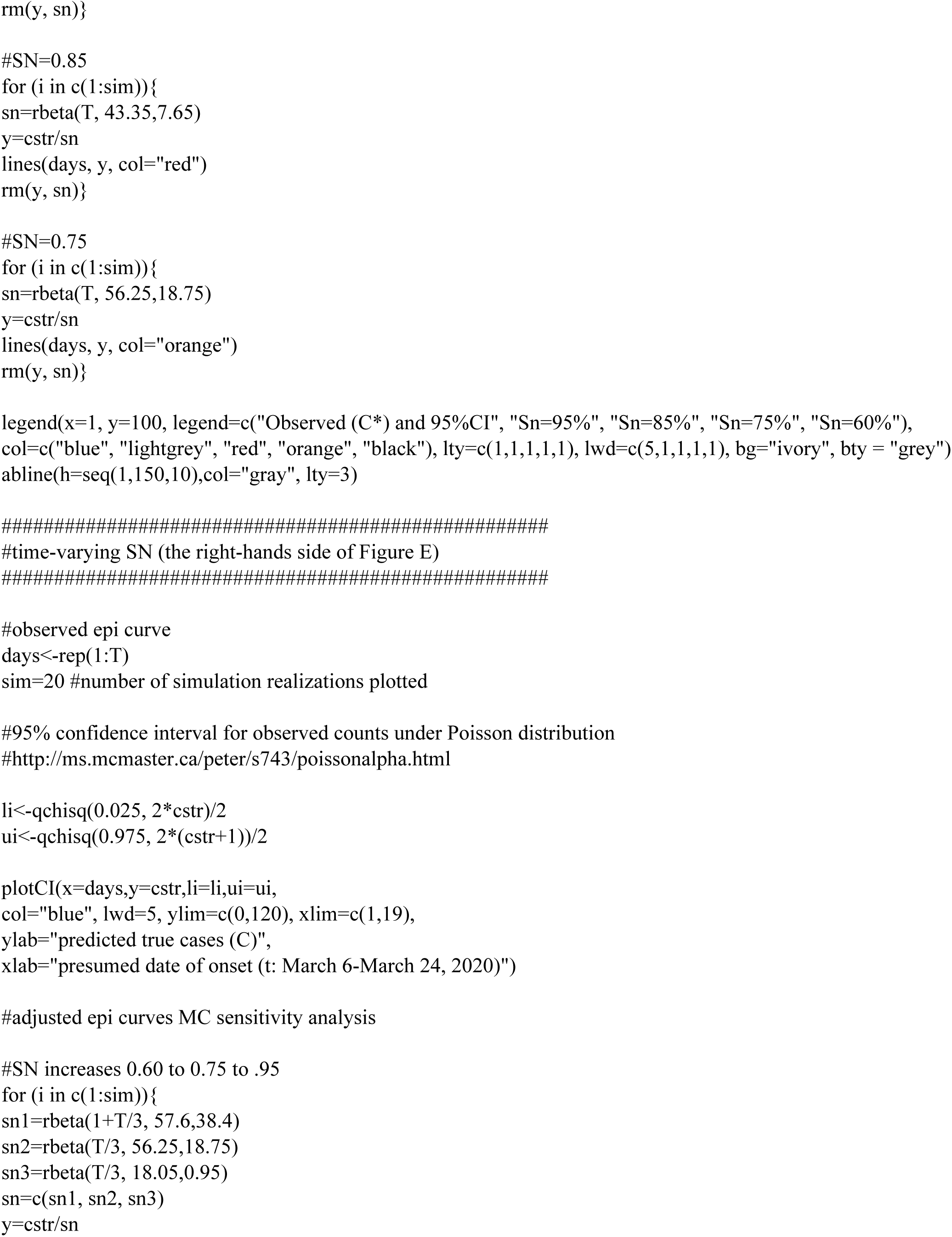
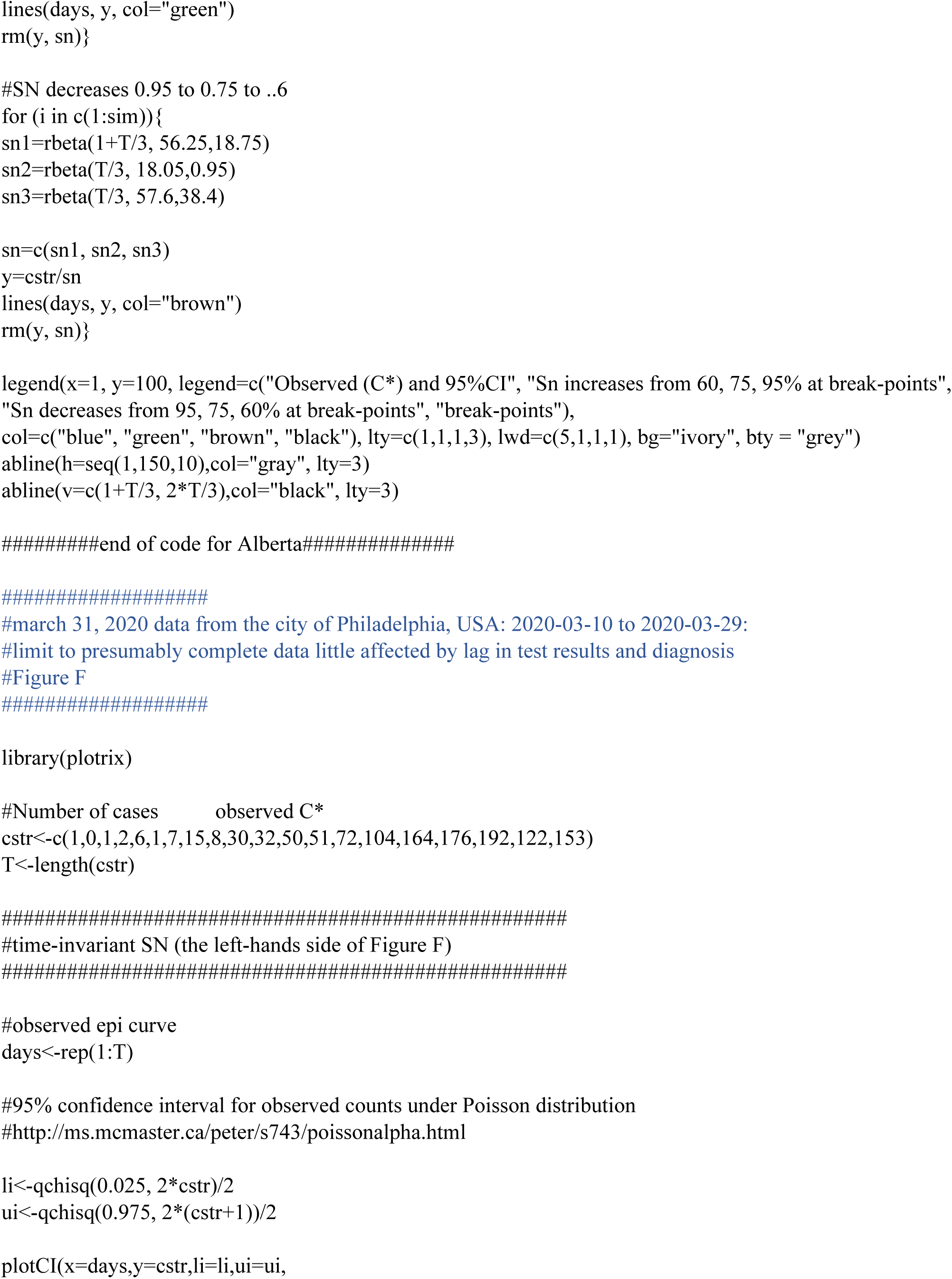
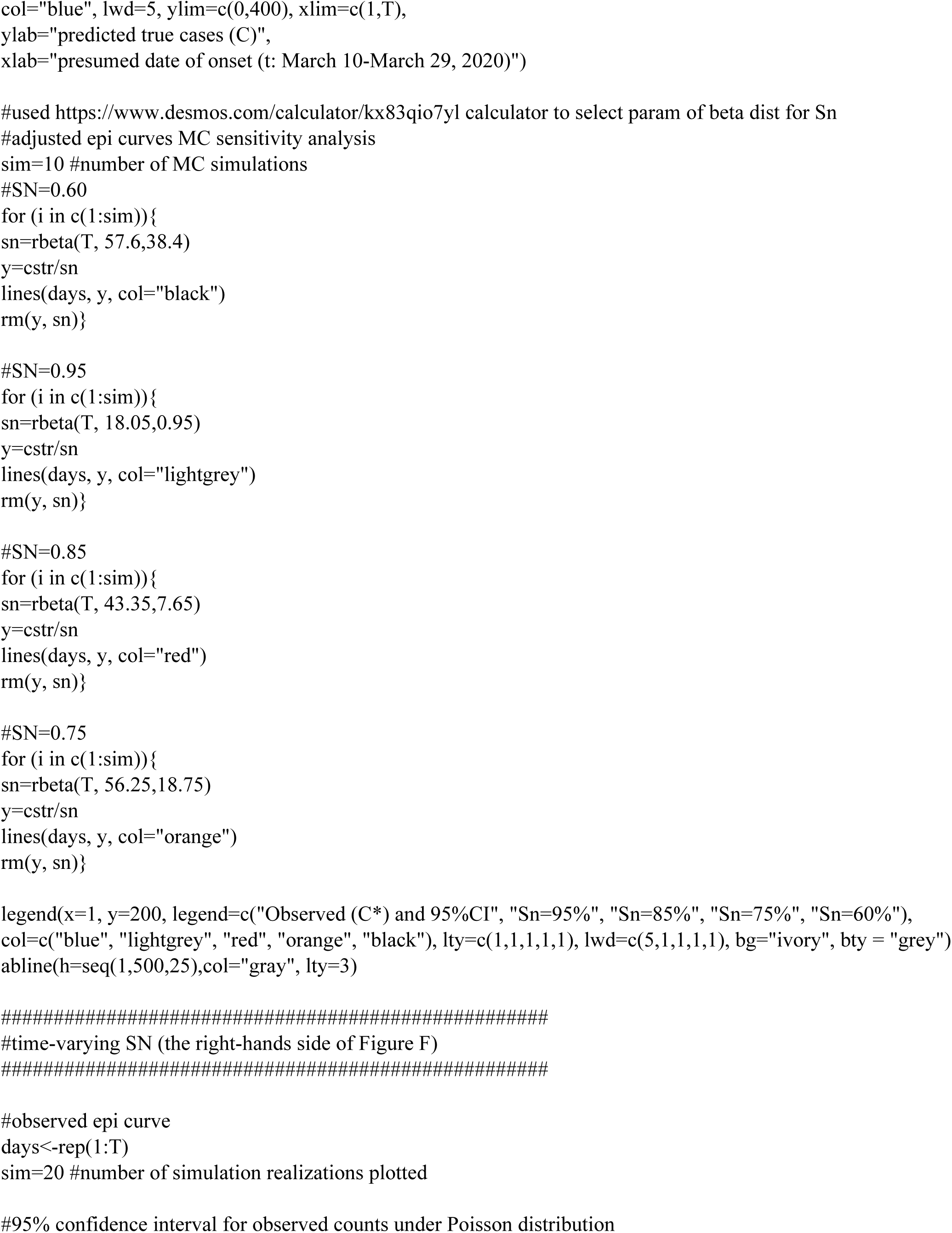
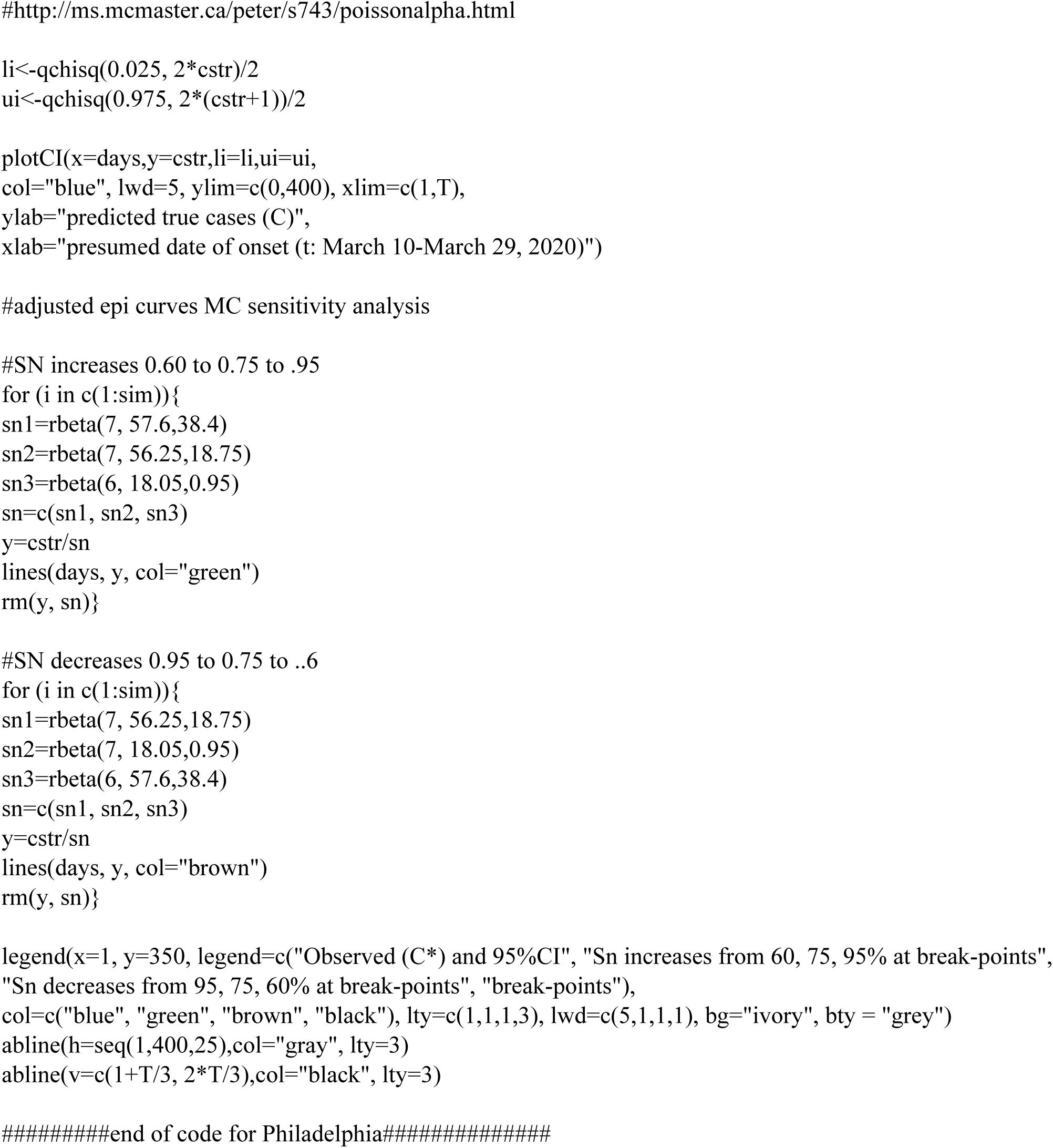

